# Prevalence and associated factors of psychological distress among patients with oral cancer in Sri Lanka

**DOI:** 10.1101/2024.06.21.24309294

**Authors:** Nadisha Ratnasekera, Irosha Perera, Pushpakumara Kandapola Arachchige, Sumeth Perera, Prasanna Jayasekara

## Abstract

**Introduction:** Oral cancer is the most common among Sri Lankan males leading to psychological distress due to its impact on the appearance and vital functions. While the negative effects of psychological distress on these patients are well known, its prevalence and associated factors remain largely unknown in Sri Lanka. This study aimed to assess the prevalence and the associated factors of psychological distress among a selected patient cohort with oral cancer in Sri Lanka.

**Methods:** A hospital-based cross-sectional study was conducted among 355 patients with oral cancer to assess the prevalence of psychological distress. A nested case-control study among 140 (per arm) patients evaluated the associated factors for psychological distress. The multivariate analysis was carried out to identify significant factors associated with psychological distress.

**Results:** The prevalence of psychological distress among patients with oral cancer was 31.0% (95% CI = 27.8%-35.3%). Being <50 years of age (Adjusted Odds Ratio (AOR)= 1.2, 95% CI= 0.7-1.7, p= 0.006), having pain (AOR (Adjusted Odds Ratio)= 44.7, 95% CI= 34-53.21, p=0.001), late stages of cancer at the diagnosis (AOR= 10.7, 95% CI= 1.07-28.78, p=0.04), being worried about basic functional disabilities (AOR= 11.4, 95% CI= 10.3-14.8, p=0.006) and the two psychological factors emerged as significant independent factors that were associated with increased risk of psychological distress among patients with oral cancer.

**Conclusion:** One-third of patients with oral cancer in the selected tertiary care hospitals suffered from psychological distress indicating its high prevalence. Our findings on associated factors of psychological distress in patients with oral cancer have contributed to the development of an intervention strategy to reduce psychological distress in this population.

- **What is already known on this topic** – Psychological distress in patients with oral cancer negatively impacts the treatment outcomes and patient’s quality of life. However, the prevalence and associated factors of psychological distress in Sri Lankan patients with oral cancer are unknown. This lack of knowledge hinders efforts to improve treatment success and patient quality of life.
- **What this study adds** – Sri Lankan patients with oral cancer experience high rates of psychological distress, with several significant contributing factors
- **How this study might affect research, practice or policy** – This study findings can guide a national program to identify and support Sri Lankan patients with oral cancer experiencing psychological distress, promoting early access to palliative care

## INTRODUCTION

The sixth leading cancer in the world is oral and pharyngeal cancer^1^. In Sri Lanka, oral cancer is the most common cancer among males, which was 15.5% of all the newly reported cancer cases in 2020. There is an increasing trend in the Age Standardized Rates of oral cancers especially in males from 1995-2020 (12.2 to 22)^2,3^.

During the diagnosis and treatment of cancer, patients undergo an array of physical and mental health problems that cause psychological distress^4,5^. Psychological distress is defined as a multifactorial, unpleasant emotional experience with physical, psychological, social, spiritual, and cultural dimensions, negatively impacting not only the quality of life of patients but their treatment outcomes as well^6^. The prevalence of psychological distress among patients with all types of cancer in the literature varies from 20% to 70%^6–11^. The limited research on psychological distress among oral cancer patients has revealed its prevalence spanning from 25% to 41%^12–15^.

Untreated psychological distress can result in low compliance with medical care, slow recovery from illness, reduction in the quality of life, poor adjustment to life after cancer treatment, high likelihood of tumor recurrence, and a low survival rate^16–21^. Hence, timely detection of psychological distress in patients with oral cancer must encompass oncological care in hospitals^22^.

Several factors are associated with the occurrence of psychological distress in patients with cancer. The National Comprehensive Cancer Network guideline on distress management (2017) has summarized the risk factors of psychological distress as: a) health-related risk factors, b) personal risk factors, c) spiritual, d) social factors, and e) other risk factors - sexual and physical abuse, substance use disorders, and other mental disorders^23^. Via a critical appraisal of the literature, it is evident that the categorization of the associated factors for psychological distress differs in each study based on a given psycho-social context. However, the basic four associated factors for psychological distress remain consistent, including socio-demographic characteristics, medical or disease-related factors, social factors, and psychological factors^22–38^.

There are minimal studies carried out in Sri Lanka to assess the prevalence of psychological distress among patients with any cancer. By using the General Health Questionnaire-12 as the measuring tool a prevalence of 69% was reported among patients with oral cancer^39^ and 30.66% among patients with breast cancer^40^. There is limited research on psychological distress among patients with cancer in Sri Lanka. Using the General Health Questionnaire-12 as an assessment tool, a prevalence of 69% was evident among patients with oral cancer^39^, while a prevalence of 30.66% was shown among patients with breast cancer^40^. To the best of the author’s knowledge, no studies have been undertaken to identify the associated factors of psychological distress among patients with oral cancer in Sri Lanka. The current study, therefore, aims to explore the prevalence and the associated factors of psychological distress among patients with oral cancer in Sri Lanka.

## MATERIALS AND METHODS

### Study design

We conducted a descriptive cross-section multi-center study with a nested case-control study. Figure 1 gives a schematic presentation of the nested case-control study.

**Figure 1:**
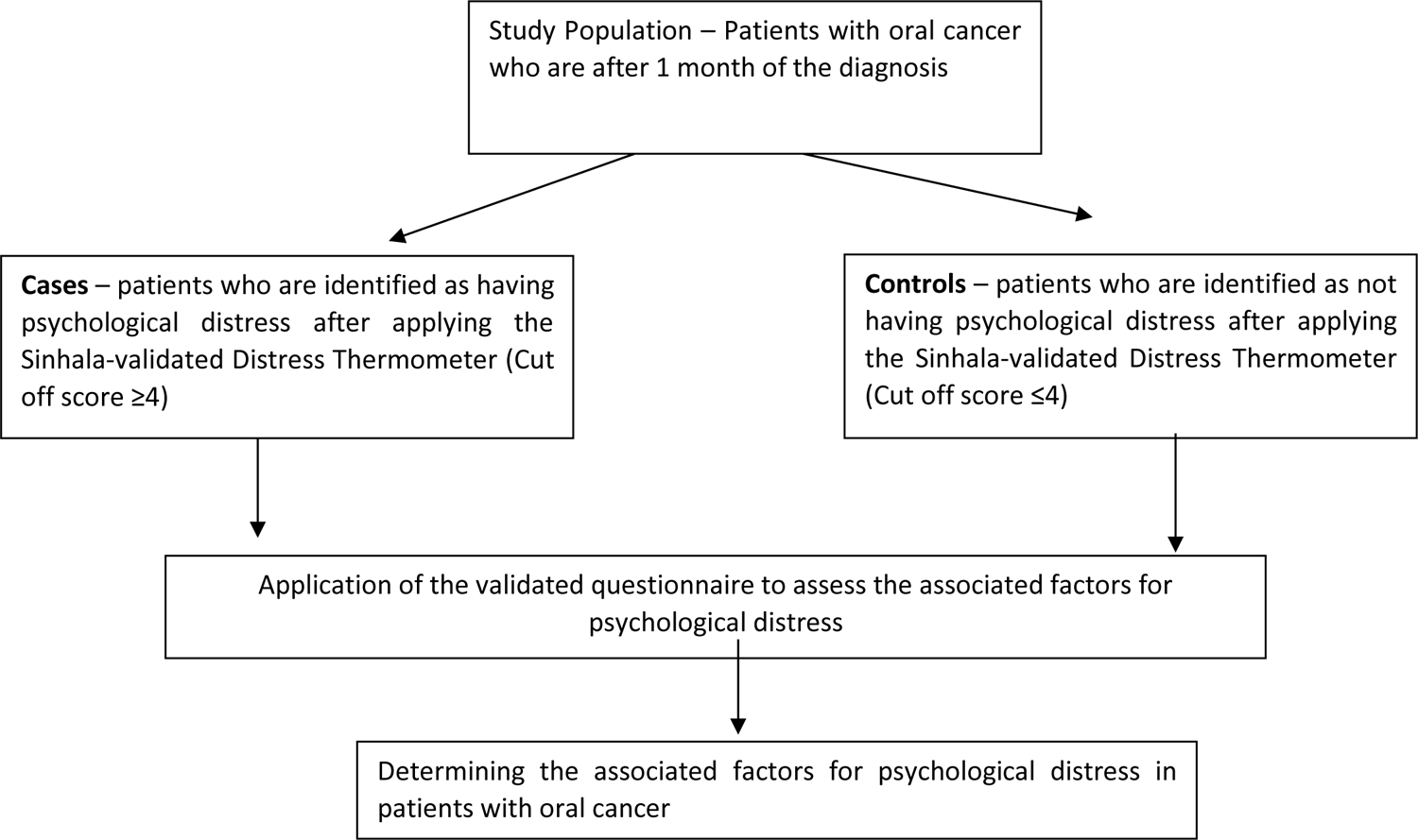
Schematic presentation of the case-control study

**Figure 2:**
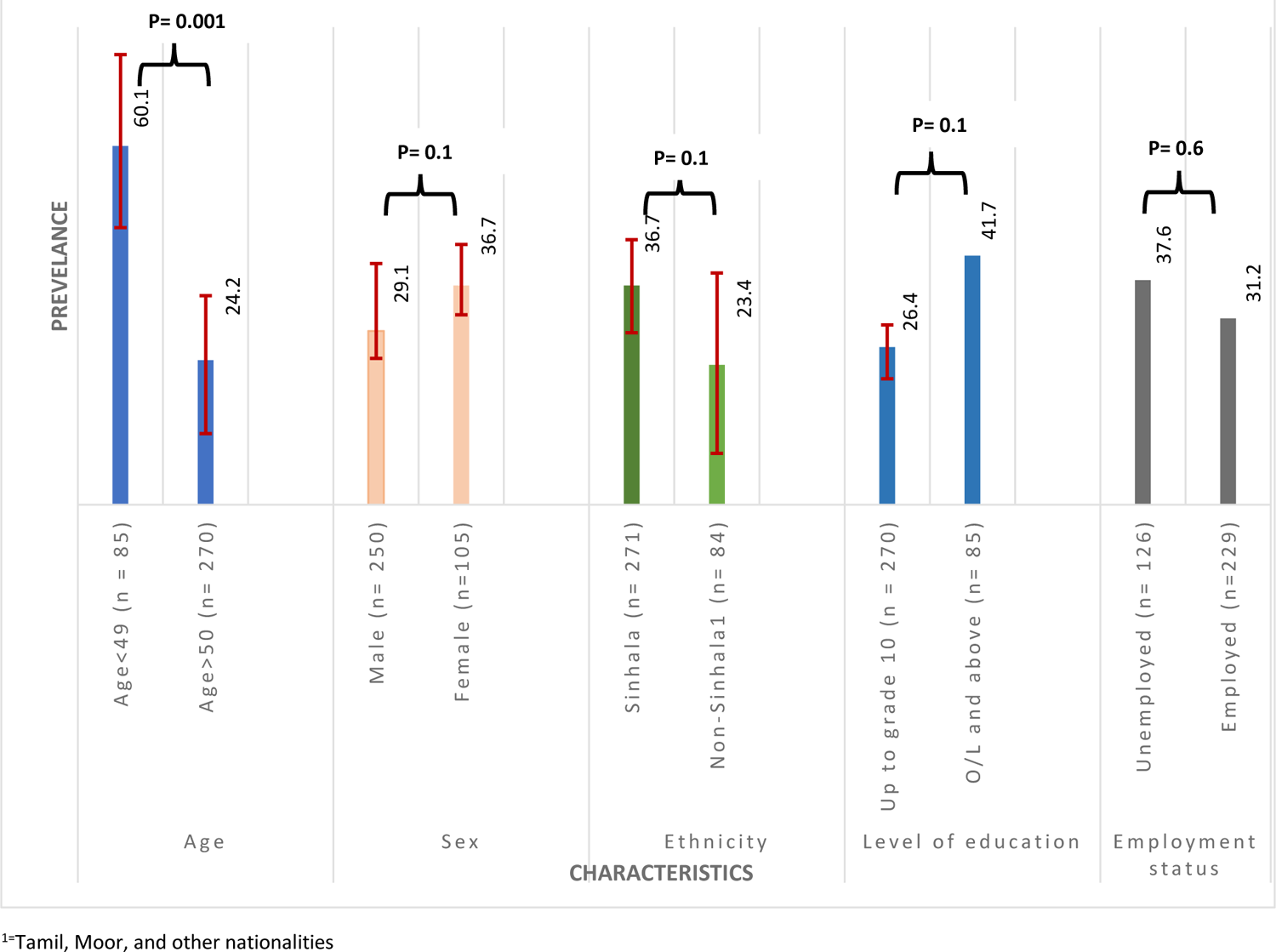
Prevalence of Psychological Distress

### Study setting and period

The study was conducted from September 2019 to January 2020, in selected tertiary care units providing oral cancer treatment in Sri Lanka (The National Dental Hospital (Teaching) Sri Lanka (NSHTSL), Colombo South (Teaching) Hospital, Apeksha Hospital - Maharagama, and Karapitiya (Teaching) Hospital).

### Study participants

The inclusion criteria were: patients who had received a definitive diagnosis of oral cancer, with clear communication about the diagnosis occurring at least one month beforehand. Additionally, the patient could be able to communicate and read well in the Sinhala language. The patients who had recurrent oral cancer, had a history of psychiatric disease, had been already treated for psychological distress at the Psychological Counseling, Spiritual and Social Development Unit, Apeksha Hospital, were receiving end-of-life palliative care, and with other severe co-morbidities were excluded from the study. All patients with oral cancer who fulfilled the eligibility criteria and gave informed consent were recruited for the study.

For the case-control study, the participants were a subset of the study units who took part in the prevalence study. The participants were divided into two groups as cases and controls. The cases included patients who were identified as having psychological distress after applying the Sinhala-validated Distress Thermometer (cut-off value ≥4)^41^. The controls included patients who were identified as **not** having psychological distress after applying the Sinhala-validated Distress thermometer (cut-off value < 4)^41^.

### Procedure

The ethical approval for this study was obtained from the Faculty of Medicine, University of Colombo, Sri Lanka (EC-18-097). Sinhala-validated Distress Thermometer and a questionnaire comprised of socio-demographic details were applied to all recruited patients. The data collection was carried out by the principal investigator with the support of a trained data collector. All the patients who were with psychological distress were recruited as cases and the first 140 patients who were without psychological distress were recruited as controls. A questionnaire to assess the associated factors of psychological distress was given to both cases and controls.

### Study size

The sample size for the prevalence study was calculated with a Z value of 1.96 at a 95% confidence interval and 0.05 precision. The anticipated prevalence of psychological distress among patients with oral cancer was 30.7%^40^, and for a 10% non-response rate, the sample size for the prevalence study was 355^41^. For the unmatched case-control study the highest sample size gained for the estimated odds ratio of the associated factors was 65 per arm after adjusting for a non-response rate of 5%^42^. However, since this case-control study was carried out on a subset of the prevalence study, all the 140 cases that were presented in the prevalence study were recruited. The cases: controls ratio was 1:1.

### Data measurement

The Sinhala-validated Distress Thermometer was used as the screening tool to identify patients with oral cancer who had psychological distress^41^. Based on a conceptual framework a multi-component, interviewer-administered questionnaire was developed to determine the associated factors of psychological distress^44^. The questionnaire was assessed for judgmental validity by 10 experts in the fields of oncology, oro-maxillo facial surgery, sociology, public health, and psychology and was pre-tested on 20 patients with oral cancer at Gampaha District General Hospital which is another tertiary care institute providing cancer care.

### Statistical methods

The SPSS version 21 was used for the data analysis. The clinically validated cut-off point of 4 was used to identify the presence of psychological distress^41^. The adjusted prevalence was presented considering the psychometric properties of the Distress Thermometer^45^.

The bivariate cross-tabulations were used to assess the associated factors for psychological distress. The strength of each variable as an associated factor for psychological distress was calculated by unadjusted odds ratio (OR) with a 95% confidence interval (Appendix I). The multivariate analysis was performed using the Logistic Regression (LR) technique to identify the Adjusted Odds Ratio (AOR) of the factors. The goodness of fit of the model was assessed with Hosmer and Lemeshow test.

## RESULTS

### Patient characteristics

The participants had a mean age of 56.72 (SD-10.57) and most of them (76.1%) were over 50 years of age and were males (70.4%). The majority (76.3%) were Sinhalese. Results indicated that 76.1% had attained an educational level up to grade 10 and 35.5% were unemployed (Table 1).

**Table 1:** Descriptive characteristics of the participants.

| **Characteristic** | **Number**<br>**(N= 355)** | **Percentage**<br>**(%)** |
| --- | --- | --- |
| **Age (Years) mean age = 56.72 (SD- 10.57)** |  |  |
| <49 | 85 | 23.9 |
| >50 | 270 | 76.1 |
| **Sex** |  |  |
| Male | 250 | 70.4 |
| Female | 105 | 29.6 |
| **Ethnicity** |  |  |
| Sinhala | 271 | 76.3 |
| Non-Sinhalese<sup>1</sup> | 84 | 23.7 |
| **Level of education** |  |  |
| Up to grade 10 | 270 | 76.1 |
| G. C.E (O/L) and above | 85 | 23.9 |
| **Occupation category** |  |  |
| Unemployed | 126 | 35.5 |
| Employed | 229 | 64.5 |

<sup>1</sup>=Tamil, Moor, and other nationalities

### Psychological distress prevalence estimations

We found that the patients with oral cancer had an overall adjusted prevalence of 31.0% (95% CI = 27.8%-35.3%) for psychological distress. In particular, the age group <49 (60.1%, 95% CI 44.8-73.8, p=0.0001) included the highest proportion among them. Moreover, considering the marital status, the married group (35.6% (95% CI = 30.3%-41.9%, p=0.01) showed a statistically significant proportion of psychological distress in the study cohort (Table 2).

### Factors associated with psychological distress

We looked at a number of parameters to find factors independently associated with psychological distress. Among them, being below 50 years of age (AOR=1.2, 95% CI: 0.7-1.7), presence of pain (AOR=44.7, 95% CI: 34.5-53.21), cancer presenting at late stages (AOR=10.7, 95% CI: 1.07-28.78), being worried about basic functional disabilities (AOR=11.4, 95% CI: 10.3-14.8), and the two psychological factors - “Other people worry about me more than I do” (AOR=5, 95% CI: 2.8-6.9) and “I feel very angry about what has happened to me”(AOR=12.1, 95% CI: 6.8-15.4) significantly increased the odds of getting psychological distress in patients with oral cancer. We applied the multivariate logistic regression model as the Hosmer-Lemeshow goodness-of-fit test (p=0.81) suggested it to be a suitable fit (Table 3).

**Table 3:** Associated factors for psychological distress of patients with oral cancer.

| Factor | Standard Error | AOR | 95% Confidence Interval |  | p-value |
| --- | --- | --- | --- | --- | --- |
| Being below 50 years of age | 1.65 | 1.2 | 0.7 | 1.7. | 0.006 |
| Presence of pain | 2.34 | 44.7 | 34.5 | 53.21 | 0.001 |
| Late stage of cancer presentation | 1.02 | 10.7 | 1.07 | 28.78 | 0.042 |
| More worried due to basic functional disabilities | 4.56 | 11.4 | 10.3 | 14.8 | 0.006 |
| “Other people worry about me more than I do” | 1.65 | 5.0 | 2.8 | 6.9 | 0.001 |
| “I feel very angry about what has happened to me” | 7.5 | 12.1 | 6.8 | 15.4 | 0.005 |

AOR – Adjusted Odd Ratio

## DISCUSSION

The current study showed 31.0% (95% CI=27.8% −35.3%) prevalence of psychological distress. We found that a number of independent variables were associated with such distress. Accordingly, certain critical factors namely those below 50 years of age, having pain, cancer presented at late stages, being worried about basic functional disabilities, and the two psychological factors (“Other people worry about me more than I do” and “I feel very angry about what has happened to me”) showed a significant association with increased distress among patients with oral cancer.

Many studies revealed that patients with all types of cancer varied in psychological distress from 20% to 70%, supporting the findings of the current prevalence study^6–11^. In contrast to the current study finding, a recent meta-analysis of the Distress Thermometer on breast cancer reported the pooled distress prevalence rate for ≥4 score in the Distress Thermometer which is the cut-off point for the current study as, 62% (95% CI 60% to 65%)^46^. This could be due to the heterogeneity of the study methods.

Despite limited research on psychological distress among patients with oral cancer using Distress Thermometer as the screening tool, a study conducted by Wang in 2018, reported a prevalence of 58.2% at ≥ 4 cut-off score on the Distress Thermometer thus replicating the cut-off score of the present study^47^. The higher prevalence than the present study could be due to variations in sample characteristics and stage of the treatment process.

There are no studies purely on the prevalence of psychological distress among patients with oral cancer in Sri Lanka. Of the available few studies among patients with other types of cancers, Mudduwa in 2011 reported a prevalence of 30.66% among patients with breast cancer which was similar to the present study finding^40^. However, in this study, the final diagnosis was made by the Consultant Psychiatrist whereas the current study used the Sinhala-validated Distress Thermometer. Another study by Weeratunga in 2016 assessed the stress levels of patients with mixed types of cancers. The screening tool used in the study was the General Health Questionnaire-12 and it was interviewer-administered which was similar to the current study. The results showed the prevalence of psychological distress in patients with oral cancer to be 69% thus indicating over two-fold increase in prevalence than the current study^40^. Such variations of the findings could be attributed to the difference in the screening tools used by the two studies and other methodological variations.

We selected a case-control study to explore the associated factors for psychological distress in patients with oral cancer as opposed to a cross-sectional study which made the evidence stronger. However, many studies use the cross-sectional study design due to obvious constraints inherent to conducting research among patients with oral cancer^48,49^. We developed the study instrument to assess the associated factors for psychological distress which was an interviewer-administered questionnaire based on the conceptual framework following a scientific process to develop new study instruments which assured the quality of data^49^. We carried out judgmental validation for the questionnaire which assured quality data collection. However, we would recommend comprehensive validation of this questionnaire in future studies.

We did not find associations with other socio-demographic factors except the younger patients had higher odds of getting psychological distress when compared with the older patients (AOR= 1.2, 95% CI 0.7%-1.7%). Grave’s cross-sectional study in 2007 supported this finding by showing that there was no association of demographic factors (sex, race, rurality of residence) with psychological distress except for age where similar to the current study younger age was related to higher levels of distress (r = −.14, p = .01)^34^. Additionally, this relationship of younger age increasing the psychological distress of patients with cancer is supported by a few other studies^27,29,47^.

A longitudinal study in eight low-and middle-income countries in Southeast Asia, a similar setting to the current study revealed a controversial finding to the current study. This study revealed that being older (>45 years) reported slightly more psychological distress than participants who were younger (Adjusted OR =1.01, 95% CI 1.01-1.02)^33^. We justify the findings of the current study by the Sri Lankan family structure and culture. Sri Lanka owns a very closely-knit family structure where the children are highly dependent on their parents as the Sri Lankan family is essentially the conjugal unit of husband, wife and dependent children. Therefore, it is rational to argue that younger and the middle-aged group of patients have a high responsibility of caring for their children and also a higher responsibility to their parents^51^.

We found no association between socioeconomic factors and psychological distress which was in contrast to the study carried out by Kimman where they found high income was associated with lesser psychological distress^33^ and the findings of Masson in 2019 where the authors revealed that patients who did not have definite employment had more stress than the people who were employed (52.6% and 26.8% respectively)^52^. We could support the current study results against these controversial findings by the fact that Sri Lanka still owns a self-sufficient and well-bonded lifestyle where the family members, extended families, neighbors, and relations see to the needs of a patient till the recovery^51^. Furthermore, Sri Lanka owns a well-established healthcare service that offers free-of-charge patient care services at the point of delivery which assures a decent financial protection for the patient^3^. Therefore, although there could be a socio-economic burden the impact on the psychological distress could be argued to be minimal.

We found that the two disease-related factors; having pain and presenting at late stages of cancer demonstrated significant association with psychological distress among patients with oral cancer. This is mainly due to pain and advanced stage with multiple complications contributing to enhanced suffering and discomfort of the oral cancer survivor which increases the levels of psychological distress. Several studies support this finding confirming pain to increase the risk of getting psychological distress in patients with cancer^29,53–55^. Kimman found the association between the cancer stage and psychological distress to be significant where the stage III patients were at a higher risk of psychological distress (Adjusted OR = 2.73, 95% CI= 2.40-3.07). Another study conducted among a group of 1496 patients with heterogeneous cancer including head and neck cancers in China revealed that the advanced stage increased the risk of psychological distress (Adjusted OR=1.85,95% CI 1.424–2.405)^33^. Therefore, the studies above supported the current study findings.

We found a significant association between ‘being worried about basic functional disabilities’ and psychological distress. This finding is in line with the majority of the studies in the literature^35,56^. Moreover, our finding of two psychological factors increased the odds of getting psychological distress in patients with oral cancer; ‘Other people worry about me more than I do’-Adjusted OR = 5; 95%CI= 2.8-6.9, ‘I feel very angry about what has happened to me’-Adjusted OR= 12; 95% CI=6.8-15.4) could be supported by few studies which revealed that worry about the loved ones around and anger about one’s own self was associated with a high risk of psychological distress in patients with cancer^35,57^.

We used the findings of this case-control study for the development of the intervention package aimed at improving the psychological distress and quality of life of patients with oral cancer which was a valid implication of the study findings^44^.

## LIMITATIONS

We acknowledge a few limitations in the study. The employment of the Distress Thermometer to classify the diseased and the non-diseased would have introduced a misclassification bias. Yet, this was minimized since the Distress Thermometer gained recognition for high diagnostic accuracy compared with the gold standard of diagnosis of Consultant Psychiatrists. Although we controlled the known confounders by multivariate analysis, we did not consider the unknown confounders and the interactions between the factors which is a limitation of the study.

We identify the exclusion of any patient who was unable to communicate in Sinhala as a limitation since Sri Lanka is a country with multi-ethnicity the ethnic and racial representation would not have been present. Hence, we recommend the necessity of replicating this study with more diverse populations.

## CONCLUSIONS AND RECOMMENDATIONS

We demonstrated that the prevalence of psychological distress among patients with oral cancer who attended the selected Public Tertiary Care Institutes in Sri Lanka was high as nearly 1/3 of patients with oral cancer suffer from psychological distress. Having pain, presenting at late stages of cancer, being worried about basic functional disabilities, and the two psychological factors - “Other people worry about me more than I do” and “I feel very angry about what has happened to me” and being below 50 years of age were significantly associated with psychological distress in patients with oral cancer.

Hence, these findings could be utilized to convince policymakers to initiate a national-level programme to screen patients with oral cancer for psychological distress and initiate sustainable programmes for the provision of early palliative care.

## Data Availability

All data produced in the present study are available upon reasonable request to the authors

## ACKNOWLEDGMENT

The authors express their gratitude to the consultants in all clinical units for their invaluable assistance in data collection.

