## Appendix I for "Prevalence and associated factors of psychological distress among patients with oral cancer in Sri Lanka"

**Appendix 1**

**The bivariate cross-tabulations to assess the associated factors for psychological distress**

*Table i: Association of Selected Sociodemographic Characteristics with Psychological Distress*

| Socio-demographic Characteristics | Total | With PD (n=140) | Without PD (n= 140) | Unadjusted OR | 95% CI | Significance |
|---|---|---|---|---|---|---|
| | | No (%) | No (%) | | | |
| **Age (N= 280)** | | | | | | |
| <50yrs | 33(100%) | 28(84.0%) | 5(16.0%) | **6.3** | **(2.0-19.0)** | **χ2 = 13.1 df = 1 p = 0.01** |
| >51yrs[a] | 247 (100%) | 112(45.3%) | 135(54.6%) | | | |
| **Sex (N=280)** | | | | | | |
| Male | 224 (100%) | 112(50.0%) | 112(50.0%) | 1 | (0.5-1.90) | χ2 = 5.44. df = 1 p = 0.95 |
| Female [a] | 56 (100%) | 28(50.0%) | 28(50.0%) | | | |
| **Ethnic Group (N=280)** | | | | | | |
| Sinhalese | 218(100%) | 116(53.3%) | 102(46.7%) | 0.5 | (0.3-1.0) | χ2 = 3.1 df = 1 p = 0.1 |
| Non-Sinhalese [a1] | 62 (100%) | 24(38.3%) | 38(61.7%) | | | |
| **Marital Status (N=280)** | | | | | | |
| Married | 235(100%) | 127(53.9%) | 108(46.1%) | 2.8 | **(1.2 – 6.2)** | **χ2 = 6.9 df = 1 p = 0.009** |
| Other categories [a 2] | 45(100%) | 13(29.4%) | 32(70.6%) | | | |
| **Education status(N=280)** | | | | | | |
| Up to grade 10 | 219(100%) | 113(51.8%) | 106(48.2%) | 0.9 | (0.4-1.4) | χ2 = 1.3 df = 1 p = 0.40 |
| O/L and above [a] | 61(100%) | 26(43.5%) | 35(56.5%) | | | |

[a] = Reference group
[1]=Tamil, Moor and other nationalities
[2]= Unmarried/ widowed/ separated or divorce

*Table ii: Association of Selected Socio-Economic Characteristics with Psychological Distress*

| Socio-economic Characteristics | Total | With PD (n=140) | Without PD (n= 140) | Unadjusted OR | 95% CI | Significance |
|---|---|---|---|---|---|---|
| | | No (%) | No (%) | | | |
| **Current employment status (N= 280)** | | | | | | |
| Un-employed | 182(100%) | 88(48.6%) | 94(51.4%) | 1.1 | (0.7-2.0) | χ2 =0.33<br>df = 1<br>P = 0.5 |
| Employed [a] | 98(100%) | 52 (52.7%) | 46 (47.3%) | | | |
| **Type of Occupation (N= 98*)** | | | | | | |
| Skilled [1] | 56(100%) | 21(38.1%) | 35(61.9%) | **4.1** | **(1.5-11.1)** | **χ2 = 8.3**<br>**df = 1**<br>**P = 0.004** |
| Non-skilled [a 2] | 42(100%) | 30(71.9%) | 12 (28.1%) | | | |
| **Is it the only source of income (N= 280)** | | | | | | |
| Yes | 74(100%) | 37(50.0%) | 37(50.0%) | 1 | (0.5-1.8) | χ2 = 0.01<br>df = 1<br>P = 0.9 |
| No [a] | 206(100%) | 103(50.0%) | 103(50.0%) | | | |
| **Faced financial difficulties due to the illness (N= 280)** | | | | | | |
| Yes | 143(100%) | 97(67.6%) | 46(32.4%) | **4.5** | **(2.5 - 7.9)** | **χ2 = 27.2**<br>**df = 1**<br>**P = 0.001** |
| No[a] | 137(100%) | 43 (31.7%) | 94(68.3%) | | | |
| **Presence of dependent children (N= 280)** | | | | | | |
| Yes | 83(100%) | 45(54.0%) | 38(46.0%) | 1.3 | (0.6-2.1) | χ2 = 0.5<br>df = 1<br>P = 0.45 |
| No[a] | 197(100%) | 95(48.3%) | 102(51.7%) | | | |
| **Marital Status of children (N= 280)** | | | | | | |
| All Married | 140(100%) | 67(48.1%) | 73(51.9%) | 1.2 | (0.7-1.9) | χ2 = 0.3<br>df = 1<br>P = 0.5 |
| All not married [a] | 140(100%) | 73(51.9%) | 67(48.1%) | | | |

[a] = Reference group
* = Out of 212 participants only 74 were employed
[1] = Professional, clerical, sales, skilled manual
[2] = Unskilled manual, market-oriented farmers and fishery workers

*Table iii: Association Between Disease-Related Factors with Psychological Distress*

| Disease Related factors | Total (%) | With PD (n=140) | Without PD (n= 140) | Unadjusted OR | 95% CI | Significance |
|---|---|---|---|---|---|---|
| | | **No (%)** | **No (%)** | | | |
| **Site of the cancer (N= 280)** | | | | | | |
| Tongue | 92(100%) | 45(48.6%) | 47(51.4%) | 1.9 | (0.6- | χ2 = 0.08 |
| Other [a1] | 188 (100%) | 95(50.7%) | 93(49.3%) | | 1.9) | df = 1 |
| | | | | | | p = 0.8 |
| **Stage of the cancer (N= 280)** | | | | | | |
| Late | 198(100%) | 115(58.0%) | 83(42.0%) | **3.1** | **(1.6-** | **χ2 = 13.1** |
| Early [a] | 82 (100%) | 25(30.6%) | 57(69.4%) | | **5.9)** | **df = 1** |
| | | | | | | **p = 0.0001** |
| **Presence of pain (N= 280)** | | | | | | |
| Yes | 117(100%) | 99 (84.3%) | 18(15.7%) | **15** | **(7.9-** | **χ2 = 72.0** |
| No [a] | 163(100%) | 41(25.2%) | 122 (74.8%) | | **32.0)** | **df = 1** |
| | | | | | | **p = 0.0001** |
| **Stage of the treatment (N= 280)** | | | | | | |
| Active treatment | 214(100%) | 132(61.7%) | 82 (38.3%) | **11.8** | **(4.8-** | **χ2 = 37.8** |
| No active treatment [a] | 66(100%) | 8(12%) | 58(88.0%) | | **29.4)** | **df = 1** |
| | | | | | | **p = 0.0001** |
| **Treated Hospital (N= 280)** | | | | | | |
| Apeksha Hospital | 87 (100%) | 45 (51.5%) | 42 (48.5) | 0.9 | (0.5- | χ2 = 0.06 |
| Other Hospitals * | 193 (100%) | 96 (49.7%) | 97 (50.3%) | | 1.7) | df = 1 |
| | | | | | | p = 0.8 |
| **Presence of co-morbidities (N= 280)** | | | | | | |
| Yes | 94(100%) | 49 (52.1%) | 45 (47.9%) | 1.13 | (0.6- | χ2 = 0.19 |
| No[a] | 186(100%) | 91(48.9%) | 95(51.1%) | | 2.0) | df = 1 |
| | | | | | | p = 0.66 |

[a] = Reference group

[1] = Palate, Buccal Mucosa, alveolus, Floor of the Mouth, lips

*Table iv: Association of Worry Related to Physical and Practical Issues with Psychological Distress*

| Worry related to physical and practical issues | Total | With PD (n=140) | Without PD (n= 140) | Unadjusted OR | 95% CI | Significance |
|---|---|---|---|---|---|---|
| | | No (%) | No (%) | | | |
| **Worry related to logistic issues (not having Lodging during treatment) (N= 280)** | | | | | | |
| Yes | 32(100%) | 27(83.3%) | 5(16.7%) | **5.9** | **(1.9-** | **χ2 = 12.0** |
| No[a] | 248(100%) | 113(45.7%) | 135(54.3%) | | **18.0)** | **df = 1** |
| | | | | | | **P = 0.001** |
| **Worry related to presence of basic functional disabilities (N= 280)** | | | | | | |
| High disability | 144(100%) | 102(70.6%) | 42(29.4%) | **6.1** | **(3.4-** | **χ2 = 38.2** |
| Low disability [a] | 136(100%) | 38(28.2%) | 98(71.8%) | | **11.1)** | **df = 1** |
| | | | | | | **P = 0.0001** |
| **Performance status (N= 280)** | | | | | | |
| Not satisfactory | 16(100%) | 15(91.7%) | 1(8.3%) | **12.1** | **(1.5–** | **χ2 = 8.8** |
| Satisfactory [a] | 264(100%) | 125(47.5%) | 139(52.5%) | | **36.1)** | **df = 1** |
| | | | | | | **P =0.003** |

[a] = Reference group

*Table v: Association of Social Factors with Psychological Distress*

| Social factors | Total | With PD (n=140) No (%) | Without PD (n= 140) No (%) | Unadjusted OR | 95% CI | Significance |
|---|---|---|---|---|---|---|
| **No of confidents (N= 280)** | | | | | | |
| 4 and below | 63(100%) | 42 (66.7%) | 21(33.3%) | **2.4** | **(1.2- 4.8)** | **χ2 = 6.9** |
| 5 and above[a] | 217 (100%) | 98(45.1%)) | 119(54.9%) | | | **df = 1** |
| | | | | | | **p = 0.009** |
| **Perceived satisfaction with confidents (N= 280)** | | | | | | |
| Not satisfied | 28(100%) | 26 (91.5%) | 2(9.5%) | **11.3** | **(2.5-9.2)** | **χ2 = 15.2** |
| Satisfied [a] | 252(100%) | 115(45.5%) | 137(54.5%) | | | **df = 1** |
| | | | | | | **p = 0.001** |
| **Presence of support groups (N= 280)** | | | | | | |
| No | 210 (100%) | 112(53.5%) | 98(46.5%) | 1.7 | (0.9-3.2) | χ2 = 3.0 |
| Yes [a] | 70(100%) | 28(39.6%) | 42(60.4%) | | | df = 1 |
| | | | | | | p = 0.02 |
| **Previous family experience of cancer (N= 280)** | | | | | | |
| Yes | 54(100%) | 32(58.5%) | 22(41.5%) | 0.6 | (0.3-13) | χ2 = 1.5 |
| No[a] | 226(100%) | 108(48.0%) | 118(52.0%) | | | df = 1 |
| | | | | | | p = 0.2 |
| **Worried about living with family members (N=280)** | | | | | | |
| yes | 125 (100%) | 82(65.3%) | 43(34.7%) | **3.1** | **(1.7-5.5)** | **χ2 = 16.0** |
| No [a] | 155 (100%) | 58(37.6%) | 97 (62.4%) | | | **df = 1** |
| | | | | | | **p = 0.0001** |
| **Worried about living with friends (N=280)** | | | | | | |
| Yes | 43(100%) | 23(54.5%) | 20(45.5%) | 1.2 | 0.6-2.6 | χ2 = 0.3 |
| No [a] | 237 (100%) | 117 (49.2%) | 120(50.8%) | | | df = 1 |
| | | | | | | p = 0.6 |

[a] = Reference group

*Table vi: Association of Factors Related to Sense of Coherence with Psychological Distress*

| **Factors Related to Sense of Coherence** | **Total** | **With PD (n=140)** | **Without PD (n= 140)** | **Unadjusted OR** | **95% CI** | **Significance** |
|---|---|---|---|---|---|---|
| | | **No (%)** | **No (%)** | | | |
| **1. 'I feel I can't do anything to cheer myself up (N=280)** | | | | | | |
| Yes | 100 (100%) | 100(100%) | 0 (0%) | **4.5** | **3.3 – 6.2** | **χ2 = 118.4** |
| No [a] | 180(100%) | 40 (22.1%) | 140 (77.9%) | | | **df = 1** |
| | | | | | | **p = 0.0001** |
| **2. I believe that my positive attitude will benefit my health (N=280)** | | | | | | |
| No [a] | 40(100%) | 39(96.7%) | 1(3.3%) | **24.3** | **20.1- 29.3** | **χ2 = 30.4** |
| Yes | 240(100%) | 101(42.3%) | 139 (57.7%) | | | **df = 1** |
| | | | | | | **p = 0.0001** |
| **3. I firmly believe that I will get better (N=280)** | | | | | | |
| Yes | 224(100%) | 99(44.1%) | 125(55.9%) | **3.6** | **1.7- 7.6** | **χ2 = 11.9** |
| No [a] | 56(100%) | 41 (73.8%) | 15(26.2%) | | | **df = 1** |
| | | | | | | **p = 0.001** |
| **4. I feel that nothing could be done to make any difference of my illness (N=280)** | | | | | | |
| Yes | 100(100%) | 100(100%) | 0(0) | **4.5** | **3.3-6.2** | **χ2 = 118.5** |
| No [a] | 180(100%) | 40(22.1%) | 140(77.9%) | | | **df = 1** |
| | | | | | | **p = 0.0001** |
| **5. I've left it all to my doctors (N=280)** | | | | | | |
| Yes | 244 (100%) | 133(54.6%) | 111(45.4%) | **5.3** | **2.1- 14.5** | **χ2 = 12.3** |
| No [a] | 36(100%) | 7(18.5%) | 29(81.5%) | | | **df = 1** |
| | | | | | | **p = 0.0001** |
| **6. I feel that life is hopeless (N=280)** | | | | | | |
| Yes | 129 (100%) | 113(87.8%) | 16(12.2%) | **33.7** | **15.5- 52.6** | **χ2 = 103.9** |
| No [a] | 151 (100%) | 26(17.5%) | 125 (82.5%) | | | **df = 1** |
| | | | | | | **p = 0.0001** |
| **7. Since my cancer diagnosis I now realize how precious life is, and I'm making the most of it (N=280)** | | | | | | |
| No | 108 (100%) | 91(84.1%) | 17(15.9%) | **13.3** | **6.5- 26.9** | **χ2 = 62.3** |

| | | | | | | |
|---|---|---|---|---|---|---|
| Yes [a] | 172(100%) | 49(28.5%) | 123(71.5%) | | | **df = 1** |
| | | | | | | **p = 0.0001** |
| **8. I have difficulty in believing that this has happened to me. I don’t really believe I have cancer (N=280)** | | | | | | |
| Yes | 135(100%) | 85(62.7%) | 50(37.3%) | **2.7** | **1.5- 4.7** | **χ2 = 12.8** |
| No [a] | 145(100%) | 55(38.2%) | 90(61.8%) | | | **df = 1** |
| | | | | | | **p = 0.0001** |
| **9. Other people worry about me more than I do (N=280)** | | | | | | |
| Yes | 236(100%) | 120(50.8%) | 116(49.2%) | **1.2** | **0.6- 2.6** | **χ2 = 0.32** |
| No [a] | 44(100%) | 20(45.5%) | 24(54.5%) | | | **df = 1** |
| | | | | | | **p = 0.06** |
| **10. I am trying to get as much as information I can about cancer (N=280)** | | | | | | |
| No | 129(100%) | 92(71.4%) | 37(28.6%) | **5.4** | **3.0-9.8** | **χ2 = 33.5** |
| Yes [a] | 151(100%) | 48(31.6%) | 103(68.4%) | | | **df = 1** |
| | | | | | | **p = 0.0001** |
| **11. I keep quite busy, so I don’t have time to think about cancer (N=280)** | | | | | | |
| No | 116(100%) | 87(75.0%) | 29(25.0%) | **6.3** | **3.4- 11.6** | **χ2 = 37.6** |
| Yes [a] | 164 (100%) | 53(32.3%) | 111(67.7%) | | | **df = 1** |
| | | | | | | **p = 0.0001** |
| **12. I feel completely lost and have no idea as to what should be done (N=280)** | | | | | | |
| Yes | 164(100%) | 128(78.2%) | 36(21.8%) | **31.5** | **14.0- 70.9** | **χ2 = 95.2** |
| No [a] | 116(100%) | 12 (10.2%) | 104(89.8%) | | | **df = 1** |
| | | | | | | **p = 0.0001** |
| **13. I am worried/ upset (N=280)** | | | | | | |
| Yes | 144(100%) | 120(83.5%) | 24(16.5%) | **29.6** | **14.0-62.4** | **χ2 = 100.6** |
| No[a] | 136 (100%) | 20 (14.5%) | 116(85.4%) | | | **df = 1** |
| | | | | | | **p = 0.0001** |

| | | | | | | |
|---|---|---|---|---|---|---|
| **14. I feel very angry about what has happened to me (N=280)** | | | | | | |
| Yes | 99(100%) | 84 (85.0%) | 15(15.0%) | **13.2** | **6.3- 27.5** | **χ2 = 57.9** |
| No [a] | 181(100%) | 55(30.7%) | 126(69.3%) | | | **df = 1** |
| | | | | | | **p = 0.0001** |
| **15. I try to fight the illness (N=280)** | | | | | | |
| No | 83(100%) | 62(74.6%) | 21(25.4%) | **4.5** | **2.3-8.6** | **χ2 = 21.7** |
| Yes [a] | 197 (100%) | 78 (39.6%) | 119(60.4%) | | | **df = 1** |
| | | | | | | **p = 0.0001** |
| **16. When I think about my life, I feel great that I am alive (N=280)** | | | | | | |
| No | 186 (100%) | 59(31.9%) | 127(68.1%) | **13.0** | **6.1- 27.8** | **χ2 = 55.0** |
| Yes [a] | 94(100%) | 81(85.9%) | 13(14.1%) | | | **df = 1** |
| | | | | | | **p = 0.0001** |

[a] = Reference group

*Table vii: Association of Other Psychological Factors with Psychological Distress*

| Factors Related to Sense of Coherence | Total | With PD (n=140) | Without PD (n= 140) | Unadjusted OR | 95% CI | Significance |
|---|---|---|---|---|---|---|
| | | No (%) | No (%) | | | |
| **1. Do you have a relaxing activity/ hobby which you get involved regularly (N=280)** | | | | | | |
| No | 116(100%) | 91(78.4%) | 25(21.6%) | **8.5** | **4.5- 16.1** | **χ2 =48.6** |
| Yes [a] | 164 (100%) | 49(29.8%) | 115(70.2%) | | | **df = 1** |
| | | | | | | **p = 0.0001** |
| **2. Addictions (N=280)** | | | | | | |
| Yes | 91(100%) | 62(68.1%) | 29(31.9%) | **3.0** | **(1.6- 5.5)** | **χ2 = 13.4** |
| No[a] | 189 (100%) | 78(41.3%) | 11/(58.7%) | | | **df = 1** |
| | | | | | | **p = 0.0001** |

[a] = Reference group

*Table viii: Association of Spiritual Factors with Psychological Distress*

| | Total | With PD (n=140) | Without PD (n= 140) | Unadjusted OR | 95% CI | Significance |
|---|---|---|---|---|---|---|
| | | No (%) | No (%) | | | |
| **Spiritual support (N=280)** | | | | | | |
| Poor | 47(100%) | 42 (88.9%) | 5(11.1%) | **11.0** | **(3.7-20.1)** | **χ2 =26.2** |
| Good [a] | 233(100%) | 98(42.0%) | 135(58.0%) | | | **df = 1** |
| | | | | | | **p = 0.0001** |

[a] = Reference group

*Table ix: Association of Existing Knowledge with Psychological Distress*

| | Total | With PD (n=140) | Without PD (n= 140) | Unadjusted OR | 95% CI | Significance |
|---|---|---|---|---|---|---|
| | | **No (%)** | **No (%)** | | | |
| **Existing knowledge (N=280)** | | | | | | |
| poor | 63(100%) | 48(77.1%) | 15(22.9%) | **4.7** | **(2.2- 9.7)** | **$\chi2 = 18.2$** |
| Good [a] | 217(100%) | 91(42.1%) | 126(57.9%) | | | **df = 1** |
| | | | | | | **p = 0.0001** |

[a] = Reference group

*Table x: Association of Satisfaction with Health Care and Psychological Distress*

| | Total | With PD (n=140) | Without PD (n= 140) | Unadjusted OR | 95% CI | Significance |
|---|---|---|---|---|---|---|
| | | **No (%)** | **No (%)** | | | |
| **Satisfaction with health care (N= 280)** | | | | | | |
| Satisfied | 272(100%) | 136(50%) | 136(50%) | 1 | (0.1- 5.0) | $\chi2 = 0.000$ |
| Not satisfied [a] | 8(100%) | 4(50%) | 4(50%) | | | df = 1 |
| | | | | | | p = 1.0 |

[a] = Reference group
